# Risk Factors for Non-Syndromic Cleft Lip and Palate : A Cross-Sectional Study

**DOI:** 10.1101/2025.05.09.25326453

**Authors:** Mohammad Mihrab Chowdhury, Miftah U Chowdhury, Nusrat Tabassum, Abu Hossain Md Moinul Ahsan, Farjana Tanni, Bijoy Krishna Das

## Abstract

**Objectives:** Orofacial clefts are among the most common congenital craniofacial anomalies, with non-syndromic forms (NSCL/P) accounting for the majority of cases. In Bangladesh, an estimated 275,000 to 300,000 individuals are affected, many lacking access to timely surgical or preventive care.

**Study Design:** This cross-sectional study examined potential contributing factors to NSCL/P among children at the pediatric surgery unit of Care Hospital in Mohammadpur, Dhaka. Data were collected over a 12-month period through structured interviews.

**Methods:** A total of 400 children with clinically confirmed NSCL/P were included. Questionnaires captured data on demographic characteristics. Univariate and multivariate analyses were conducted to identify associations risk factors.

**Results:** Both analyses revealed that young maternal age, low maternal education and delay fatherhood associated with increased risk.

**Conclusions:** These findings highlight the role of socioeconomic and demographic factors in the occurrence of NSCL/P. The results have implications for health professionals and public health, specifically, early identification of at-risk groups.

## 1. Introduction

Cleft lip and/or palate (CL/P) affect thousands of newborns globally each year ^[1, 2]^. These malformations result from incomplete fusion of facial structures during early embryonic development and may present as cleft lip (CL), cleft palate (CP), or both ^[3, 4, 5]^. CL/P is typically classified into syndromic (SCL/P) and non-syndromic (NSCL/P) forms ^[6]^. NSCL/P occurs without associated anomalies and constitutes approximately 55% of all cases ^[7, 8, 9, 10]^.

Despite advances, genetic and regional mechanisms underlying CL/P are not fully understood ^[11, 12, 13]^. The etiology of CL/P remains complex and multifactorial ^[14]^. Both genetic predispositions and environmental exposures—including maternal smoking, alcohol use, poor nutrition, infections, and medication use during pregnancy—have been implicated ^[15, 16, 17, 18]^. The relative influence of these risk factors can vary widely by population due to socioeconomic, cultural, and dietary differences ^[19, 20, 21]^. In low- and middle-income countries, factors like malnutrition, environmental toxins, and inadequate prenatal care may further elevate risk ^[22]^.

NSCL/P can significantly affect a child’s development, leading to feeding difficulties, malnutrition, speech delays, and psychosocial challenges. Infants often experience difficulties in breastfeeding due to impaired sucking reflexes, contributing to malnutrition and developmental delays ^[23, 24, 25]^. As affected children grow, they may encounter speech impairments and social stigma. ^[26]^.These burdens extend to families and health systems, particularly where specialized care is limited.

Global studies have demonstrated that the prevalence and type of cleft conditions vary across populations. For example, Asian and Native American groups exhibit higher rates (around 1 in 500 live births), while African populations show lower incidence rates ^[16, 11, 27, 28]^. According to the Global Burden of Disease (GBD) study, oceania reports the highest rates of CP, whereas Sub-Saharan Africa records the lowest ^[29, 30]^. Similarly, data from EUROCAT registries in Europe show marked variation, with Finland and Malta reporting the highest CP rates and Portugal and Romania the lowest ^[5, 28]^. Within South and Southeast Asia, regional disparities are also pronounced. For instance, in Pakistan, Punjab accounts for the majority of CL/P cases ^[31]^, and studies in India (e.g., Chennai and Gujarat) confirm a higher male prevalence and variation in cleft types ^[32, 33]^. Nepal reports an annual CL/P prevalence of 1.64 per 1,000 live births, with a higher incidence in females^[34]^. Comparable trends have been noted in the Palestinian Territories, Taiwan, the Philippines, Thailand, Singapore, and Malaysia—where CL/P is more common among males and often associated with low birth weight ^[35, 32]^.

Due to its prevalence, NSCL/P presents a more pressing public health concern than its syndromic counterpart. The absence of other congenital anomalies in NSCL/P makes it diagnostically and etiologically challenging. This complexity underscores the importance of identifying underlying factors—both genetic and environmental. Moreover, treatment for NSCL/P typically involves multidisciplinary care, including prenatal screening, surgical intervention, and long-term support for speech development, feeding, and psychosocial well-being. Such comprehensive treatment places a considerable burden on healthcare systems, especially in low-resource settings. In Bangladesh, approximately 275,000 to 300,000 people live with cleft conditions ^[36]^. Despite this substantial burden, there remains a lack of population-specific research exploring the prevalence and risk factors associated with NSCL/P.

This study aims to address this gap by identifying maternal, paternal, family, environmental, and birth-related risk factors potentially associated with NSCL/P in children treated at a pediatric surgical hospital in Dhaka. Data were collected through structured interviews during clinical visits. the responses were then analyzed to examine how these factors may contribute to the occurrence of NSCL/P. Understanding these associations is essential for informing public health interventions and optimizing access to surgical care. This will ultimately reduce the burden of NSCL/P in resource-limited settings such as Bangladesh.

## 2. Methods

### 2.1. Study design

This cross-sectional study evaluated five categories of potential risk factors associated with NSCL/P: maternal factors, paternal and family factors, environmental exposures, birth-related characteristics, and socio-demographic variables. Maternal factors included prenatal conditions such as diabetes, anaemia, folic acid supplementation, tobacco use (including smoking), medication intake, and exposure to environmental toxins during the first trimester. Paternal and family factors focused on exposure to household smoking by parents and other family members. Environmental factors assessed maternal exposure to insecticides and radiation. Birth-related variables included mode of delivery, birth order, and blood group. Socio-demographic characteristics encompassed the child’s age, and gender, parental age, religious affiliation, birth defects and education, family history, and household economic status. The outcome variable was the presence of NSCL/P confirmed by clinical diagnosis. A conceptual framework outlining the relationship between variables is presented in Figure 1. Data collection was conducted over a 12-month period (January 1 to December 31, 2023) at the pediatric surgery unit of Care Hospital in Dhaka, Bangladesh. Ethical approval was granted by the National Institute of Preventive and Social Medicine (NIPSOM), and written informed consent was obtained from the participants.

**Figure 1:**
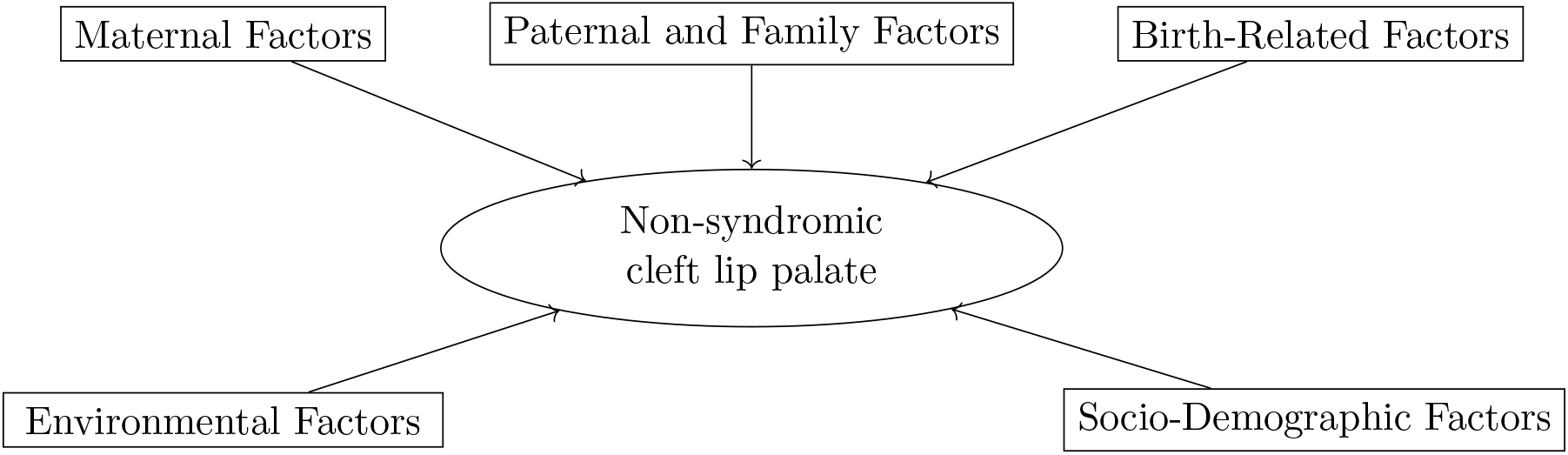
Computational Framework

### 2.2. Analysis

All collected data were checked for completeness and consistency. We also de-identified the data to maintain participant confidentiality prior to analysis. Descriptive statistics were used to summarize the distribution of NSCL/P subtypes across the study population. Associations between catergorical variables and cleft subtypes were initially assessed using chi-square test. To further investigate the relationship between potential risk factors and NSCL/P, three regression models were employed: logistic regression (LR), Poisson regression with robust standard errors(PR), and log-binomial models (LB). LR was used to estimate adjusted odds ratios (OR) for risk factors. PR was selected for its suitability in studies with smaller sample sizes ^[37]^. LB was applied directly to calculate relative risks (RRS). The use of multiple regression approaches allowerd for comparison and validation of results, ensuring robustness and consistency of the findings ^[38]^. All statistical analyses were perfomed using SPSS (version 23.0) and Python (version 3.4.1).

## 3. Result

### 3.1. Descriptive Analysis

Among the 400 children diagnosed with NSCL/P, the most common presentation was combined cleft lip and palate (NSCLP) (67.0% cases), followed by isolated cleft palate (NSCP) (9.0%), and isolated cleft lip (NSCL) (14.0%) (Table 1). Laterality patterns varied across cleft types. Among childred with NSCL (n=56), left-sided clefts were more prevalent (51.8%), followed by right-sided clefts (39.3%), with bilateral clefts being the least frequent (Table B.2). In the NSCLP subgroup (n=268), left-sided clefts were present in 39.9% of cases, right-sided clefts in 31.0%, and bilateral clefts in 32.7% (Table B.3). For children with NSCP, the majority (91.9%) had involvement of both the hard and soft palate, indicating a predominance of extensive clefting within this subgroup (Table B.4). The predominance of left-sided clefts is consistent with prior findings^[26, 32]^

**Table 1:** Distribution of respondents according to their type of NSCL/P.

| Distribution of NSCL/P | Frequency (f) | Percentage (%) |
| --- | --- | --- |
| NSCL only | 56 | 14.0 |
| NSCLP | 268 | 67.0 |
| NSCP only | 76 | 19.0 |
| **Total** | **400** | **100.0** |

### 3.2. Study Population

The cohort comprised 251 males (62.75%) and 149 females (37.25%). First-born children represented the largest group (41.5%), followed by second born children (33.0%). Cesarean delivery was more common (58.25%) was more common than vaginal delivery (41.50%). The majority of mothers were aged between 20–34 years (72.25%), and more than half had completed education up to Class 10. Similarly, 52.75% of fathers had completed education up to Class 10, while only 21.00% had attained higher education.

Several variables exhibited high proportions of missing data, including family history (95.75%), blood relation (79.25%), blood group (75.00%). In addition to missingness, some variables were highly skewed, limiting their utility in inferential analysis. For instance, maternal smoking was reported as absent in 99.50% of cases, maternal diabetes in 80.25%, and maternal insecticide exposure in 86.75%. Given these limitations, the following variables were excluded from further analysis: religion, child age, maternal tobacco use, maternal smoking, maternal diabetes, maternal insecticide exposure, family history, blood groups, and maternal anemia (Table B.1).

### 3.3. Analysis of Factors

We applied multiple regression models and chi-square tests to assess the association between individual variables and cleft types while controlling for potential confounders.

#### Univariate

Univarite chi-square analysis (Table 2) revealed that birth order and paternal education were significantly associated with NSCLP. Paternal education also appeared as a significant factor for NSCL. In contrast, gender and monthly income were significantly associated with NSCP.

**Table 2:** Adjusted ORs for chi square statistic with different types of NSCL/P.

| NSCL/P | Significant | Chi-square | P-Value | Degress of Freedom |
| --- | --- | --- | --- | --- |
| NSCL | Paternal Education | 7.427866 | 0.024381 | 2 |
| NSCLP | Birth Order | 9.093302 | 0.028076 | 3 |
|  | Paternal Education | 10.016229 | 0.006683 | 2 |
| NSCP | Gender | 10.326872 | 0.001311 | 1 |
|  | Monthly Income | 6.103831 | 0.047268 | 2 |

To refine these associations, we employed three regression models: LR, PR and LB. All three models (Table 3, Table C.5, Table C.6) produced consistent results. In LR model, (Table 3),maternal age between 20 and 35 years acted as a protective factor, while lower maternal education and higher paternal education (Hons and above) were associated with significantly increased odds of cleft occurrence.

**Table 3:** Adjusted ORs for univariate logistic regression associated with different types of NSCL/P with p *<* 0.05. Other model results are available in appendix (Table C.5, Table C.6)

| NSCL/P | Significant | Reference | Adjusted OR | 95% CI | P Value |
| --- | --- | --- | --- | --- | --- |
| NSCL | Maternal Age 2 | 1 | 0.5 | [0.33, 0.93] | 0.0283 |
|  | Maternal Education 2 | 1 | 2.04 | [1.08, 3.85] | 0.028 |
|  | Paternal Education 3 | 1 | 2.51 | [1.27, 4.93] | 0.008 |
| NSCLP | Paternal Education 2 | 1 | 0.55 | [0.33,0.91] | 0.019 |
|  | Paternal Education 3 | 1 | 0.47 | [0.27, 0.799] | 0.005 |
| NSCP | Female | Male | 2.342 | [0.344, 1.358] | < 0.01 |
|  | Monthly Income 2 | Monthly Income 1 | 0.6468 | [0.3102, 1.3486] | 0.2451 |
|  | Monthly Income 3 | Monthly Income 1 | 1.814 | [0.9628, 3.4176] | 0.0654 |

For NSCLP, higher paternal education showed a protective effect across models. In NSCP cases, females had significantly higher odds. Additionally, income levels between $20,000 and $29,999 appeared protective compared to lower income groups, whereas income above $30,000 was paradoxically associated with increased risk, which may be related delayed fatherhood ^[39]^. For NSCP, second-born child had higher odds in both PR and LB models. Paternal education group 2 emerged as risk factor in PR, while maternal age between 20 and 35 years was protective in LB. When analyzing laterality in NSCLP (unilateral vs. bilateral), chi-square tests did not reveal significant associations; however, all three regression models identified birth order as a contributing factor. Specifically, second-born children had increased risk for unilateral NSCLP and decreased odds for bilateral NSCLP when compared to first-born children.

#### Multivariate

Variables identified as significant in the univariate analysis were further examined using multivariate models (Table 4, Table D.7, Table D.8). For NSCP, maternal age between 20 and 35 years consistently emerged as a protective factor across all models. In contrast, paternal education group 3 (Honors and above) was associated with an increased risk of NSCP. For NSCLP, both moderate (SSC to HSC) and higher paternal education (Honors and above), as well as, being a second child, were protective in both the LR and PR models. In the LB model, higher paternal education (group 3) also showed a protective association. Among children with NSCP, female was found to be a strong and consistent risk factor across all models. Additionally, the PR model identified maternal age group 3 (*>* 35 years) and paternal education group 2 (SSC to HSC) as risk factors for NSCP.

**Table 4:**
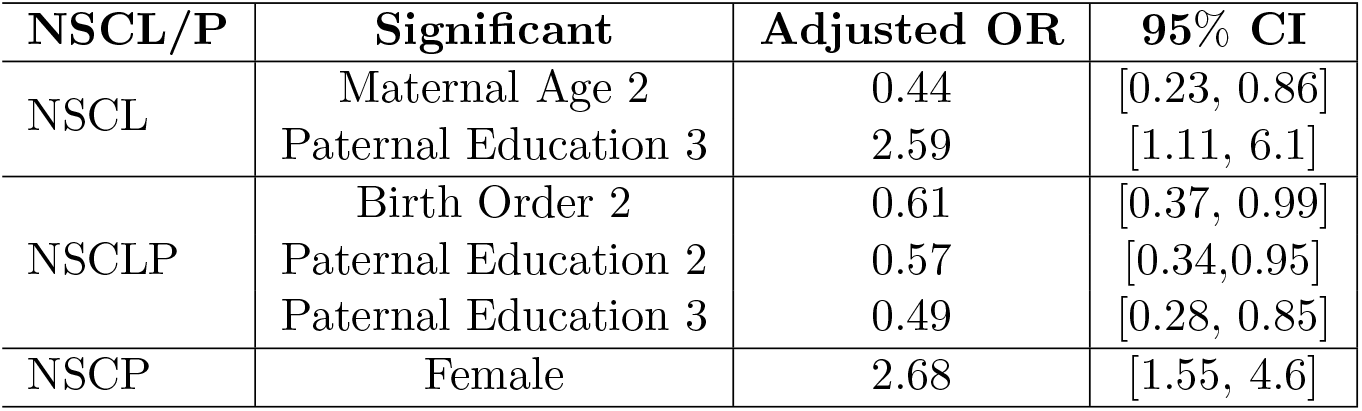
Adjusted ORs for multivariate logistic regression associated with different types of NSCL/P with p *<* 0.05.

| NSCL/P | Significant | Adjusted OR | 95% CI |
| --- | --- | --- | --- |
| NSCL | Maternal Age 2 | 0.44 | [0.23, 0.86] |
|  | Paternal Education 3 | 2.59 | [1.11, 6.1] |
| NSCLP | Birth Order 2 | 0.61 | [0.37, 0.99] |
|  | Paternal Education 2 | 0.57 | [0.34, 0.95] |
|  | Paternal Education 3 | 0.49 | [0.28, 0.85] |
| NSCP | Female | 2.68 | [1.55, 4.6] |

## 4. Discussion

This study highlights the multifactorial nature of NSCL/P and underscores the interplay of demographic, socioeconomic, and biological influences in a pediatric population in Bangladesh. While the etiology of NSCL/P has a strong genetic component, our findings reinforce the significant role of modifiable factors such as parental education, maternal age, gender, and birth order - each varying in influence across cleft subtypes.

Consistent with previous research, female gender was significantly associated with isolated cleft palate (NSCP). This essentially suggests potential hormonal or gender-linked developmental influences during embryogenesis. ^[40, 1]^. A significant proportion of parents had only secondary education. This reflects broader disparities in health awareness and access to preventive care, which are known barriers to maternal health and early intervention in low- and middle-income countries (LMICs) ^[41, 42]^. Although anemia and folic acid supplementation were commonly reported among mothers, their associations with cleft risk were unclear—possibly due to variations in dietary quality, supplement timing, dosage, or adherence ^[43, 44, 45]^.

Univariate analysis identified that birth order and paternal education were significantly associated with NSCL and NSCLP, while females and lower household incomes were associated with NSCP. These findings support the notion that distinct cleft subtypes may have different etiological profiles. Multivariate regression models revealed that maternal age between 20 and 34 years was a consistent protective factor, particularly for NSCL. Higher paternal education (honors and above) showed a dual effect—acting as a risk factor in NSCL but as a protective factor in NSCLP. This pattern may reflect greater healthcare-seeking behavior among more educated families or be confounded by age-related paternal risk factors ^[39]^. Moreover, the association of paternal and maternal age as risk factors may differ depending on whether parental ages are examined independently or in combination ^[46]^. For NSCLP, medium-to-higher paternal education and being second-born were related to reduced odds. This may suggest either intrauterine environmental changes between pregnancies or evolving maternal health behaviors over time. Being female is associated with nearly a twofold increase in the risk of NSCP compared to males. Additionally, income between $20,000 and $29,999 appeared protective for NSCP. Higher income levels (*>* $30,000) were associated with increased risk which may be related to delayed fatherhood ^[39]^. These associations suggest that cleft subtypes differ in their sensitivity to both biological and parental factors. ^[47, 48, 5, 46]^.

From a surgical standpoint, these findings are critical for early risk stratification and resource planning. Early cleft repair, ideally within the first year of life, significantly improves speech development, feeding, and psychosocial integration ^[49]^. However, delayed diagnosis—especially in underserved areas—may hinder timely surgical care. Identifying high-risk groups based on parental education, maternal age, and birth order can facilitate earlier intervention in cleft care programs. As this study was conducted in a Smile Train-affiliated pediatric surgical center, it provides valuable insight into children accessing cleft care. However, it also highlights the need for future community-based studies to capture data on untreated or undiagnosed cases.

Nonetheless, the study has limitations. Even though our study provided critical insights into children receiving cleft care at a low-resource setting, it is based on a single Smile Train-affiliated pediatric surgical unit. Some variables could not be considered due to missingness or skewness. Moreover, the observed protective effect of maternal age (20–35 years) in one model suggests further investigation into family and reproductive factors. These limitations highlight opportunities to build on this work through larger, multi-center studies that incorporate prospective data collection, genetic profiling, and more precise tracking of environmental exposures.

To summarize, this study contributes to a growing body of evidence supporting the multifactorial etiology of NSCL/P. While genetic predisposition plays a central role, our results show that many risk factors are socially and environmentally mediated. Strengthening maternal education, delaying early pregnancies, improving prenatal counselling, and ensuring timely referral to surgical care can collectively reduce the burden of NSCL/P. Integrating these insights into national cleft management policies and outreach programmes will be essential for advancing surgical outcomes across LMICs.

## 5. Conclusions

This cross-sectional study, conducted at the pediatric surgery unit of Care Hospital in Dhaka, identified key demographic and socioeconomic risk factors for NSCL/P. Lower parental education, younger maternal age (*≤*19 years), and higher birth order were associated with increased risk. Maternal age between 20–34 years and higher paternal education appeared to be protective. Gender-related differences were also noted, particularly in cleft palate cases. These findings highlight the importance of addressing modifiable factors through public health strategies, including delaying early pregnancies, improving maternal education, and expanding access to prenatal care. Community-based programs that raise awareness and support maternal health may contribute to reducing cleft incidence. Future research should include larger, population-based samples and explore genetic–environmental interactions to support comprehensive prevention efforts.

## Data Availability

All data produced in the present study are available upon reasonable request to the authors

## 6. Ethics Statement

Ethical approval for this study was obtained from the Ethical Review Committee of the National Institute of Preventive and Social Medicine (NIPSOM), Bangladesh (Approval No. NIPSOM/IRB/2023/06). Written informed consent was obtained from all participants after providing information about the study’s purpose, procedures, and confidentiality measures. Participation was voluntary, and no biological samples or invasive procedures were involved.

## 8. Acknowledgement

All authors declare no competing interests. They have no financial or personal relationships that could have inappropriately influenced this work. All authors had access to the data, approved the final manuscript, and agreed to be accountable for the integrity and accuracy of the study. We would like to thank, Dr. Kripalini Roy, Orthodontic Surgeon, New Life Hospital Ltd, for helping with the data collection.

## Appendix

### A. Questionnaire

After preliminary observation and a review of the relevant literature, a semi-structured questionnaire was developed as the primary research instrument. Initially prepared in English, the questionnaire was translated into Bangla and subsequently back-translated into English by an independent third researcher to ensure the accuracy of the translation. The questionnaire was designed using selected variables based on the specific objectives of the study. A pretest was conducted with 20 respondents at Islami Bank Hospital, Motijheel, Dhaka, and necessary revisions were made based on the feedback received. Both the English and Bangla versions of the finalized questionnaire used for data collection will be provided upon request.

The first section gathered baseline socio-demographic information, including age, sex, and religion (categorized as Islam, Hinduism, Buddhism, or Christianity). The second section captured clinical details such as the position of the cleft lip and/or palate, blood group (ABO and Rh status), birth order based on live births, and mode of delivery (normal vaginal delivery or cesarean section). The third section collected parental information, including age, educational qualifications (primary, secondary, higher secondary, graduation, post-graduation), average monthly family income (in Bangladeshi currency), history of smoking or tobacco use, history of miscarriage, disabled child births, and presence of blood relation between parents. The fourth section focused on maternal health during the first trimester, including history of infection, fever, anemia, diabetes mellitus, use of medications (iron, folic acid, and vitamins), and exposure to insecticides, mosquito coils, or x-rays.

### B. Demographics

**Table B.1:** Summary of Variables with Frequency, Proportion, Unique Values, and Missing Values.

| Variable | Category | Frequency | Proportion (%) |
| --- | --- | --- | --- |
| Sex | Male | 251 | 62.75 |
|  | Female | 149 | 37.25 |
| Blood group | Missing | 300 | 75.00 |
|  | B+ | 33 | 8.25 |
|  | A+ | 30 | 7.50 |
|  | O+ | 26 | 6.50 |
|  | O- | 3 | 0.75 |
|  | A- | 3 | 0.75 |
|  | ab+ | 3 | 0.75 |
|  | ab- | 2 | 0.50 |
| Birth order | First | 166 | 41.50 |
|  | Second | 132 | 33.00 |
|  | Third | 85 | 21.25 |
|  | Fourth or More | 17 | 4.25 |
| Mode of delivery | Cesarian Delivery | 233 | 58.25 |
|  | Normal Delivery | 166 | 41.50 |
| Maternal Age | $\leq 19$ years | 88 | 22.00 |
|  | 20 years - 34 years | 289 | 72.25 |
| | $\geq 35$ years | 23 | 5.75 |
| Maternal Education | Upto Class 10 | 217 | 54.25 |
|  | Class 10 to HSC | 113 | 28.25 |
|  | Honours and Higher | 70 | 17.50 |
| Paternal Age | $\leq 19$ years | 4 | 1.00 |
|  | 20 years - 34 years | 288 | 68.75 |
| | $\geq 35$ years | 121 | 30.25 |
| Paternal Education | Upto Class 10 | 211 | 52.75 |
|  | SSC to HSC | 105 | 26.25 |
|  | Honours and Higher | 84 | 21.00 |
| Blood Relation | No | 357 | 89.25 |
|  | Yes | 43 | 10.75 |
| Maternal Smoking | No | 398 | 99.50 |
|  | Yes | 2 | 0.50 |
| Maternal Tobacco | No | 377 | 94.25 |
|  | Yes | 23 | 5.75 |
| Paternal Smoking | No | 293 | 73.25 |
|  | Yes | 107 | 26.75 |
| Family Member Smoking | No | 333 | 83.25 |
|  | Yes | 67 | 16.75 |
| Abortion | No | 286 | 71.50 |
|  | Yes | 114 | 28.50 |
| Birth Defect | No | 373 | 93.25 |
|  | Yes | 27 | 6.75 |
| Maternal Illness | No | 301 | 75.25 |
|  | Yes | 99 | 24.75 |
| Maternal Fever | No | 241 | 60.25 |
|  | Yes | 158 | 39.50 |
|  | Missing | 1 | 0.25 |
| Maternal Diabetes | No | 321 | 80.25 |
|  | Missing | 65 | 16.25 |
|  | Yes | 14 | 3.50 |
| Maternal Anemia | No | 242 | 60.50 |
|  | Yes | 98 | 24.50 |
|  | Missing | 60 | 15.00 |
| Maternal Folic Acid | Yes | 277 | 69.25 |
|  | No | 118 | 29.50 |
|  | Missing | 5 | 1.25 |
| Maternal Medicine | No | 226 | 56.50 |
|  | Yes | 170 | 42.50 |
|  | Missing | 4 | 1.00 |
| Maternal Insecticide | No | 347 | 86.75 |
|  | Yes | 49 | 12.25 |
|  | Missing | 4 | 1.00 |
| Maternal Coil | No | 217 | 54.25 |
|  | Yes | 178 | 44.50 |
|  | Missing | 5 | 1.25 |
| Maternal Radiation | No | 254 | 63.50 |
|  | Yes | 139 | 34.75 |
|  | Missing | 7 | 1.75 |
| Family History | No | 328 | 82.00 |
|  | Missing | 55 | 13.75 |
|  | Yes | 17 | 4.25 |
| Monthly Income | Missing | 6 | 1.5 |
| | < \$20000 | 256 | 64.0 |
| | \$20000 - \$29999 | 77 | 19.25 |
| | ≤ \$30000 | 61 | 15.25 |

**Table B.2:** Distribution of cleft lip laterality among individuals with NSCL only.

| Cleft Lip Laterality | Frequency (f) | Percentage (%) |
| --- | --- | --- |
| Left-sided cleft lip | 29 | 51.8 |
| Right-sided cleft lip | 22 | 39.3 |
| Bilateral cleft lip | 5 | 8.9 |
| **Total** | **56** | **100.0** |

**Table B.3:** Distribution of NSCLP among respondents.

| Both Cleft Lip and Palate | Frequency (f) | Percentage (%) |
| --- | --- | --- |
| Left side cleft | 107 | 39.9 |
| Right-sided cleft | 83 | 31.0 |
| bilateral clefts | 78 | 32.7 |
| **Total** | **268** | **100.0** |

**Table B.4:** Distribution of NSCP position among respondents.

| Position of Cleft Palate | Frequency (f) | Percentage (%) |
| --- | --- | --- |
| Both hard and soft palate | 316 | 91.9 |
| Only soft palate | 28 | 8.1 |
| **Total** | **344** | **100.0** |

### C. Univariate

**Table C.5:** Adjusted odds ratios for univariate Poisson Regression Results with Robust Variance different types of NSCL/P with p *<* 0.05.

| NSCL/P | Significant Variables | Reference Variable | Rate Ratio | 95% CI | P Value |
| --- | --- | --- | --- | --- | --- |
| NSCL | Maternal age 2 | 1 | .56 | [0.34, 0.93] | .02 |
|  | Maternal education 2 | 1 | 1.83 | [1.07, 3.15] | 0.02 |
|  | Paternal education 3 | 1 | 2.17 | [1.24, 3.8] | 0.006 |
| NSCLP | Paternal education 2 | 1 | 0.82 | [0.69, 0.98] | 0.03 |
|  | Paternal education 3 | 1 | 0.77 | [0.63, 0.95] | 0.012 |
| NSCP | Female | Male | 1.97 | [1.31, 2.95] | < 0.01 |
|  | Birth Order 2 | 1 | 1.58 | [1.009, 2.49] | 0.04 |
|  | Paternal Education Group 2 | 1 | 1.58 | [0.9628, 3.4176] | 0.04 |

**Table C.6:**
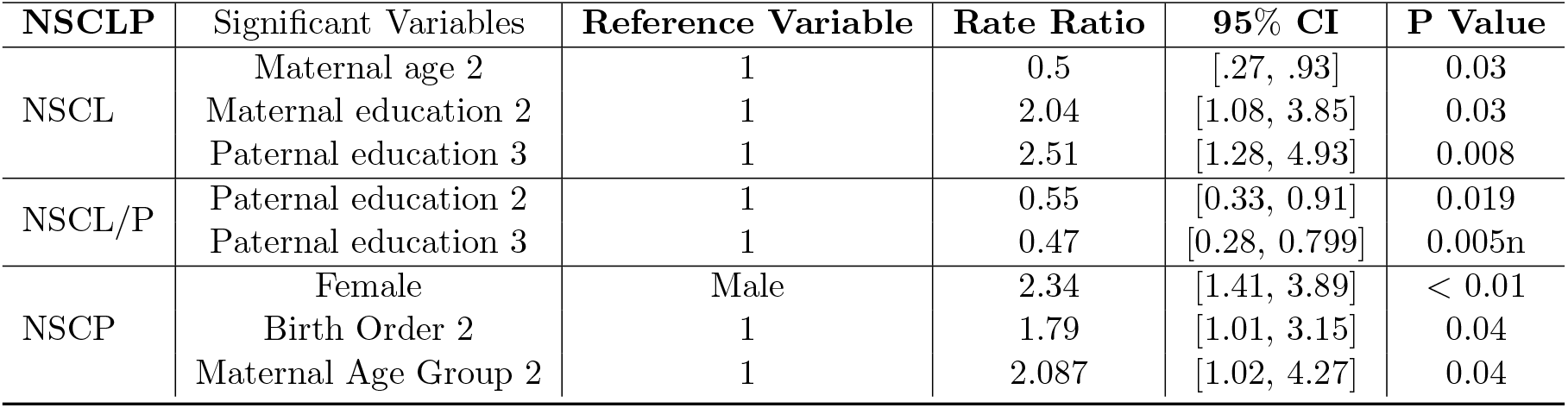
Adjusted odds ratios for univariate Log-Binomial Results with Robust Variance different types of NSCL/P with p *<* 0.05.

### D. Multivariate

**Table D.7:** Adjusted odds ratios for multivariate Poisson Regression Results with Robust Variance different types of NSCL/P with p *<* 0.05.

| NSCL/P | Significant Variables | Rate Ratio | 95% CI |
| --- | --- | --- | --- |
| NSCL | Maternal age 2 | .58 | [0.31, 0.86] |
|  | Paternal education 3 | 2.17 | [1.15, 4.1] |
| NSCLP | Birth Order 2 | 0.84 | [0.7, 1] |
|  | Paternal Education 2 | 0.84 | [0.7, 0.99] |
|  | Paternal Education 3 | 0.78 | [0.64, 0.96] |
| NSCP | Female | 2.11 | [1.4, 3.2] |
|  | Maternal Age 3 | 2.6 | [1.03, 6.3] |
|  | Paternal Education Group 2 | 1.58 | [1, 2.5] |

**Table D.8:** Adjusted odds ratios for multivariate Log-Binomial Results with Robust Variance different types of NSCL/P with p *<* 0.05.

| NSCL/P | Significant Variables | Rate Ratio | 95% CI |
| --- | --- | --- | --- |
| NSCL | Maternal age 2 | 0.5 | [.3, .83] |
|  | Paternal education 3 | 2.19 | [1.128, 4.25] |
| NSCLP | Paternal education 3 | 0.8 | [0.65, 0.96] |
| NSCP | Female | 2.05 | [1.38, 3.1] |

